# Cost-benefit analysis of deceased donor organ transplantation: an economic evaluation

**DOI:** 10.64898/2026.03.18.26348687

**Authors:** James Riley, Barnaby Roberts, Bilal Rashid, Laura Barton, Matthew Wellberry Smith, Ruth Sutcliffe, Emma Billingham, Stephen Pettit, Gareth Jones, Andrew J. Fisher, Jasvir Parmar, Sern Lim, Rommel Ravanan, Derek Manas

**Author notes:** Correspondence to Barnaby Roberts, Department of Health and Social Care, 39 Victoria Street, London SW1H 0EU.

## Abstract

**Background:** Each year around 4,500 people in the UK receive an organ transplant. These surgeries can be life changing and life extending for patients but are also associated with significant costs for the health service. However, by reducing the need for other expensive interventions involved in non-transplant care for organ failure, such as dialysis, some of these costs may be recovered.

**Methods:** We assessed the lifetime costs and benefits associated with transplantation focussing on deceased donor adult transplants for kidneys, livers, hearts, and lungs. We incorporated costs of organ retrieval, surgery, post-transplant secondary care, and medications, as well as impacts on quality and duration of life. These were compared to the cost of managing patients with end-stage organ failure for whom no transplant occurs.

**Results:** Kidney transplants were found to be cost saving with lifetime costs approximately £220,000 lower than alternative treatment. Heart transplants and liver transplants were found to be more cost-effective than thresholds used by NICE for new medicines at approximately £17,000 to £18,000 per quality adjusted life year gained. Lung transplants were the least cost-effective organ transplant with a cost per quality adjusted life year gained of over £50,000.

**Conclusions:** Although transplants can be costly, not providing a transplant to a patient who needs one also brings significant costs. Kidney transplants can save the health system money by reducing the need for dialysis. Increasing the number of kidney’s available for transplant could save the NHS money whilst saving and improving lives.

## Introduction

Despite a great amount of innovation and research within organ donation and transplantation the UK continues to face a shortage of organs for transplant. As of July 2025, 7,949 people were on the active organ transplant waiting list. Between 1 April 2024 and 31 March 2025, 463 people died waiting for a transplant, and a further 911 patients were removed from the transplant list due to deteriorating health [1].

Modelling by Kidney Research UK suggests that by 2033 the annual demand for kidney transplants could be 11,665 but with capacity to perform just 3,615 [2]. The unmet need is likely to result in significant dialysis costs to the NHS which the same report estimates at £34,000 per patient per year.

To address these challenges, the Department of Health and Social Care in England established the Organ Utilisation Group (OUG) which made twelve recommendations in their report focused on maximising the potential for organ utilisation [3].

Improving organ utilisation is now a central focus within transplantation, with significant resources dedicated to meeting demand and enhancing patient outcomes. NHS Blood and Transplant have obtained initial investment to explore the establishment of a national Assessment and Recovery Centre (ARC) service, a key recommendation of the OUG report that could enable an additional 750 transplants per year.

To support this transformation effort, an economic evaluation of transplantation services is essential. This analysis builds on and significantly adapts work by analysts from the Department of Health and Social Care (DHSC) for a 2018 impact assessment to analyse the value of moving to an opt-out system of organ donation [4]. Many studies have assessed the cost-effectiveness of different types of transplant such as [5-8], but we are not aware of any other recent assessment of the economic impact of transplantation across several types of solid organs.

The scope of this assessment is lifetime healthcare costs and benefits associated with deceased donor transplants of kidneys, liver, lungs, or heart to adult patients in the UK.

## Methods

The modelling estimates future costs and quality-adjusted life years (QALYs) associated with two pathways, one where transplantation occurs, and one where it does not (“transplant” path and “no transplant” path). For each group, annual costs and QALYs are considered for the remainder of the patient’s life. Costs are discounted at 3.5% and health benefits at 1.5% per year in line with the HM Treasury’s Green Book [9]. Discounting is used to allow costs and benefits that occur on different time spans to be compared on a common “present value” basis.

The scope of the modelling has been limited to health impacts to the patient and healthcare costs faced by the National Health Service (NHS). Wider impacts such as employment, impacts on family members, or impacts on the environment have not been included.

For heart transplants we conducted separate analysis for patients on the three different waiting lists: non-urgent, urgent, and super-urgent. For lung transplants we just consider non-urgent and urgent lists as the sample size for the super-urgent waiting list was too small to provide meaningful results.

Parameter values and sources are listed in **Error! Reference source not found**.. Health related quality of life utility values are taken from [10 & 11].

Costs for transplant surgery are taken from the national cost collection for the NHS [12]. As these figures vary significantly from year to year, averages of financial years 2022-23, 2023-24, and 2024-25 were used.

Annual post-transplant secondary care costs were derived by linking Hospital Episodes Statistics data with national cost collection values via Healthcare Resource Group codes. From year four onwards the annual cost was held flat at the lowest of the first three years cost.

Medication costs were based on information provided by clinicians for their estimate of doses given to a typical patient. For kidney, liver, and heart recipients, the majority of the total medication cost is from tacrolimus or alternatives. Medication costs for lung transplant recipients are higher in the first year due primarily to antifungals such as caspofungin and antibiotics such as colomycin. Costs are based on British National Formulary list prices [13]. This could be an overestimate of the true costs paid as price reductions are often achieved due to the purchasing power of centralised immunosuppression prescribing. In our later sensitivity analysis, we include a scenario with a 25% reduction in medication costs that shows a slight increase in cost-effectiveness.

A cost of £74,580 per donation, and £29,050 per transplant was included for organ retrieval. This was assumed based on information provided directly by NHS Blood and Transplant (NHSBT) and accounts for operational costs and overheads but excludes costs of transformation projects. These figures include costs from non-proceeding donors.

For the counterfactual, to estimate the treatment costs when a transplant is not carried out, the costs in the year before transplant of a patient who did receive a transplant are used. These costs were derived from Hospital Episodes Statistics (HES) and the national cost collection in the same way as the post-transplant secondary care costs. This approach was chosen to estimate waiting list costs whilst using the same set of patients as in the treatment scenario for a fairer comparison. Two exceptions to this approach were made.

For patients on the non-urgent heart transplant waiting list, the costs from the year preceding the year preceding transplant were used. HES does not list which waiting list a patient is on and so costs in the year before transplant are likely to be higher than is typical for patients on the non-urgent list who may remain non-urgent for several years. By using costs from the year preceding the year preceding transplant, we obtain a steadier estimate of costs without transplant. Patients on the non-urgent waiting list are assumed to eventually become urgent and then super-urgent before dying in the event that no transplant is carried out. At this point the costs associated with the urgent and super-urgent lists are applied.

For patients on the super-urgent heart transplant waiting list the daily unit cost of a heart critical care bed was used.

As HES does not include dialysis, dialysis costs were added for kidney patients in the counterfactual using figures from the national cost collection for the NHS. The proportion of patients on haemodialysis and peritoneal dialysis was based on data from the 2025 UK Renal Registry 27th Annual Report [14]. Transport costs of attending haemodialysis were estimated at £4,110 per patient per year based on uplifting an estimate from [15]. The cost of high-cost drugs not covered by the dialysis tariff, namely erythropoietin, were included based on estimates from the Organ donation for transplantation costing report uplifted to £5,117 per patient in 2024/25 prices [16]. It was assumed that 80% of those on dialysis would receive erythropoietin.

Post-transplant survival is based on 1-year and 5-year from the NHSBT organ specific reports 2024-25 and median survival data provided by NHSBT [17]. Survival was modelled to be piecewise linear between these points. Beyond the median we have continued to assume a linear decline in survival but have also applied a mortality factor based on the 2025 ONS lifetables to account for all-cause mortality on the basis that patient age as well as graft age will become significant factors for survival [18].

Survival without a transplant was based on judgement from clinicians informed by data on waiting list survival provided by NHSBT. The waiting list survival data was not used directly as patients who remain on the waiting list until death were not thought to be a fair reflection of all patients who enter the waiting list. Most patients are either transplanted or removed from the waiting list before death. Clinicians were asked to provide high and low estimates of average survival from the point at which a transplant would typically occur, if no transplant took place. Survival curves are shown in Figure 1.

**Figure 1.**
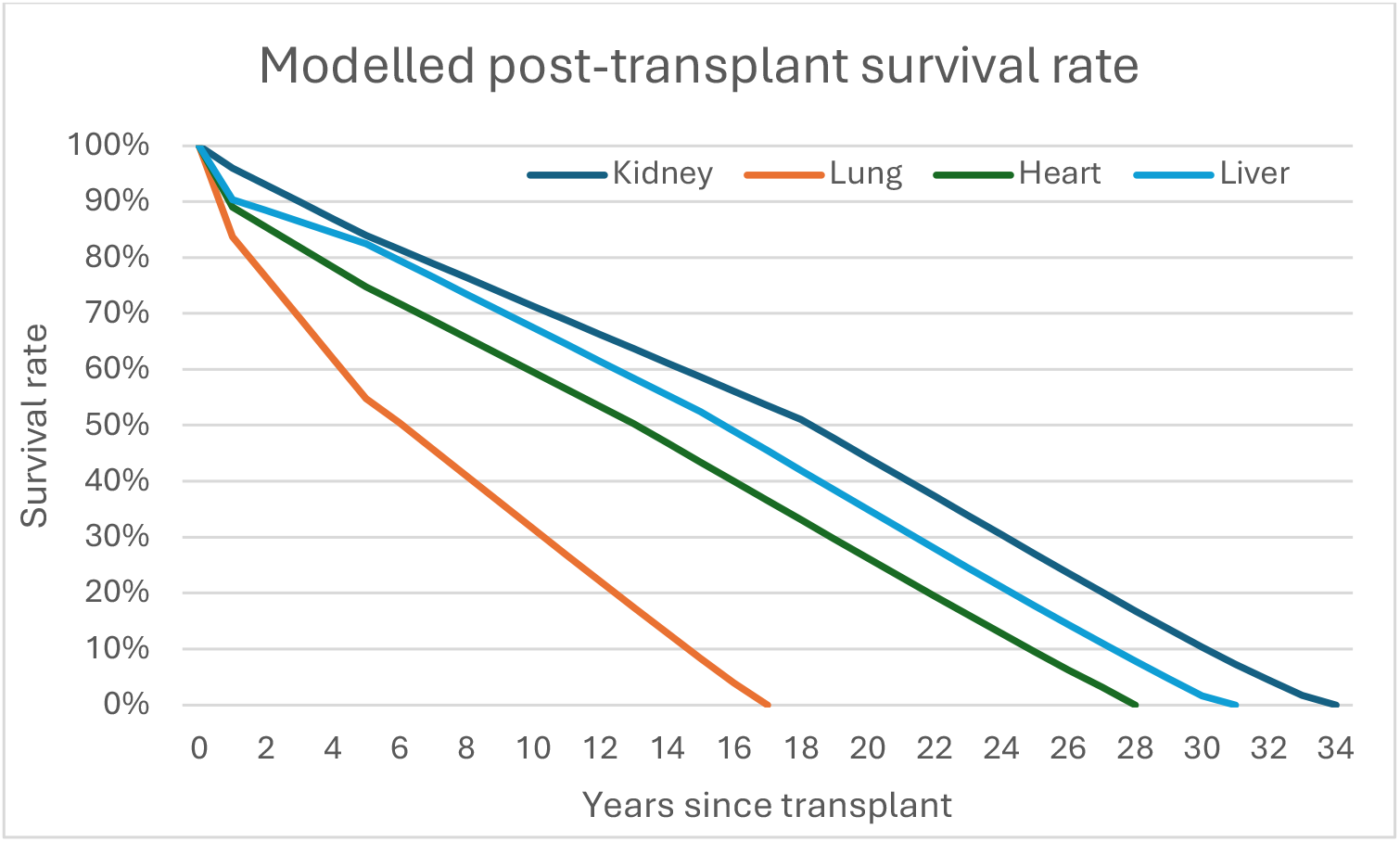
Modelled post-transplant survival rates.

The median survival on the waiting list of kidney patients is 10.8 years according to NHSBT statistics with a median waiting time of 1.3 years. Based on this a median survival rate of 10 years was assumed in the counterfactual with a range of 5 to 15 years.

For patients on the non-urgent lung waiting list median survival is 2.7 years with a median waiting time of 1.5 years. Clinicians agreed on a central estimate of 1.75 years survival in the counterfactual (range 0.5 to 3.0 years). For urgent lung patients, clinicians agreed an average counterfactual survival of two months.

For patients on the non-urgent heart waiting list the median survival is 5.8 years. However, many people move from the non-urgent list to the urgent list so the average survival if no organ become available is likely to be lower than this. Clinicians agreed a central assumption of four years survival in the counterfactual with a range of two to six years.

Of patients on the urgent heart waiting list who have not been transplanted six months after being listed, about 60% have either died or being removed from the list. Clinicians agreed on a range of three to twelve months survival in the counterfactual with a central estimate of 7.5 months. For those on the super-urgent list survival in the counterfactual is assumed to be 3.5 months in our central scenario with a range of one to six months.

No NHSBT estimate of median survival on the waiting list was available for liver patients. However, of those who have not been transplanted two years after being listed, 35% have died and a further 25% have been removed from the list. Clinicians supported using a central assumption of two years survival in the counterfactual with a range of one to three years.

The model is highly sensitive to some of the input parameters and so sensitivity analysis was performed by varying each of the key input parameters by plus-or-minus 25%. Survival without transplant is likely the most uncertain parameter so for the sensitivity analysis this was varied by plus-or-minus 50% which closely resembles the ranges provided by clinicians.

Modelling was conducted in MS excel and is available on request. No patient and public involvement was used in this study.

## Results

### Error! Reference source not found

shows the headline central estimates of organ transplant cost-effectiveness. Kidney transplants are found to provide significant cost savings primarily through averting the need for dialysis, with an incremental cost reduction of £223,619 for every kidney transplant performed, over the course of the patient’s life. With 2,299 adult kidney transplants from deceased donors taking place in the UK in 2024-25 [17] this equates to over half a billion pounds in savings over the lifetime of these patients.

Lung transplants are found to be the least cost-effective at over £50k per QALY gained. This is largely due to post-transplant survival being worse than for other organ transplants and because survival without a transplant is poor and therefore has lower costs.

Heart and liver transplants are found to be similarly cost-effective at around £16-18k per QALY gained.

Considering the average number of kidney, lung, heart, and liver transplants per donor gives an estimated saving of £203k per donor and a gain of 18.1 QALYs.

For the purposes of improving organ utilisation through measures such as ARCs, it is appropriate to exclude retrieval costs as these costs would have been encountered even if the organ were deemed unsuitable for use. Outputs that exclude retrieval costs are shown in Table 3.

**Table 1.** parameters and sources.

|  | Kidney | Lung | Heart | Liver | Source |
| --- | --- | --- | --- | --- | --- |
| **Health related quality of life utilities** |  |  |  |  | Values for kidney, liver, and heart are from [10]. Values for lung are from [11]. |
| Post transplant | 0.76 | 0.89 | 0.75 | 0.78 |  |
| Waiting list | 0.56 | 0.65 | 0.25 | 0.42 |  |
| **Costs** |  |  |  |  |  |
| Transplant surgery cost | £19,376 | £72,986 | £90,055 | £36,339 | Activity weighted average unit costs from HRG codes. |
| Annual secondary care cost w/o transplant |  |  |  |  | Derived using HES data linked to HRG codes for the year before transplant. Except in the case of non-urgent heart waiting list where the year before the year before transplant was used, and the super-urgent heart waiting list where a heart critical care cost of £2098 per day was used. For kidney, patients' dialysis costs are added along with costs of transport to hospital dialysis, and high-cost drugs not covered by the dialysis tariff. |
| Standard/non-urgent waiting list | £57,017 | £22,045 | £14,167 | £34,982 |  |
| Urgent waiting list | NA | £22,045 | £28,710 | NA |  |
| Super urgent waiting list | NA | NA | £765,610 | NA |  |
| Post-transplant secondary care costs |  |  |  |  |  |
| Year 1 (excluding surgery) | £27,379 | £63,197 | £94,584 | £33,009 | Derived using HES data linked to HRG codes. |
| Year 2 | £11,723 | £19,496 | £15,356 | £11,585 |  |
| Year 3 | £10,936 | £21,485 | £15,844 | £10,141 |  |
| Year 4+ annual | £10,936 | £19,496 | £15,356 | £10,141 |  |
| Medication costs |  |  |  |  | Schedules and some cost information provided by clinicians with additional cost information from the British National Formulary. |
| Year 1 | £5,926 | £12,250 | £2,884 | £3,331 |  |
| Year 2+ annual | £3,409 | £2,860 | £2,883 | £3,241 |  |
| Organ retrieval cost | £74,580 per donor, £29,050 per transplant |  |  |  | NHSBT personal communication. |
| Organs utilised per donor | 1.69 | 0.22 | 0.14 | 0.60 | NHSBT activity report [1]. |
| One-year patient survival | 96% | 84% | 89% | 90% | NHSBT organ specific reports [17]. |
| Five-year patient survival | 84% | 55% | 75% | 83% | NHSBT organ specific reports [17]. |
| Median patient survival | 18.4 | 6.1 | 13.1 | 15.8 | Provided by NHSBT. |
| Annual morbidity after five years (excluding all cause mortality) | 2.5% | 4.4% | 3.1% | 3.0% | Based on 5-year and median survival. |
| Average age of recipient | 53 | 57 | 49 | 49 | NHSBT activity report and organ specific reports [1,17]. |
| Expected survival w/o transplant (years) |  |  |  |  | Estimates were guided by information available in the NHSBT activity reports on waiting list survival but ultimately based on judgements from clinicians. |
| Standard/non-urgent waiting list |  |  |  |  |  |
| High | 15 | 3 | 6 | 3 |  |
| Central | 10 | 1.75 | 4 | 2 |  |
| Low | 5 | 0.5 | 2 | 1 |  |
| Urgent waiting list |  |  |  |  |  |
| High | NA | 0.25 | 1.00 | NA |  |
| Central | NA | 0.17 | 0.63 | NA |  |
| Low | NA | 0.08 | 0.25 | NA |  |
| Super urgent waiting list |  |  |  |  |  |
| High | NA | NA | 0.50 | NA |  |
| Central | NA | NA | 0.29 | NA |  |
| Low | NA | NA | 0.08 | NA |  |
| Discount rate (health impacts) | 1.50% |  |  |  | In line with HM Treasury's Green Book approach [9]. |
| Discount rate (costs) | 3.50% |  |  |  |  |

**Table 2.** Headline cost-effectiveness results per transplant (with discounting)

|  | Kidney | Lung non-urgent | Lung urgent | Heart non-urgent | Heart urgent | Heart super-urgent | Liver |
| --- | --- | --- | --- | --- | --- | --- | --- |
| Transplant costs | £243,324 | £284,106 | £284,106 | £372,878 | £372,878 | £372,878 | £237,345 |
| Counterfactual costs | £466,943 | £38,366 | £3,674 | £244,213 | £232,873 | £223,303 | £67,923 |
| Transplant QALYs | 11.19 | 5.74 | 5.74 | 8.55 | 8.55 | 8.55 | 10.26 |
| Counterfactual QALYs | 5.12 | 1.15 | 0.11 | 0.95 | 0.16 | 0.07 | 0.83 |
| Incremental costs | -£223,619 | £245,740 | £280,432 | £128,665 | £140,005 | £149,575 | £169,421 |
| Incremental QALYs | 6.07 | 4.59 | 5.63 | 7.59 | 8.39 | 8.47 | 9.43 |
| Cost per QALY gained | -£36,856 | £53,541 | £49,818 | £16,944 | £16,685 | £17,650 | £17,963 |

**Table 3.** Cost-per-QALY with retrieval costs excluded.

|  | Kidney | Lung non-urgent | Lung urgent | Heart non-urgent | Heart urgent | Heart super-urgent | Liver |
| --- | --- | --- | --- | --- | --- | --- | --- |
| Cost per QALY gained | -£41,644 | £47,212 | £44,657 | £13,118 | £13,223 | £14,222 | £14,883 |

### Sensitivity analysis

There is significant uncertainty on some of these estimates as highlighted by the sensitivity analysis below.

### Error! Reference source not found

Table 4 shows the estimated cost-per-QALY-gained for each organ type under different parameter adjustments. In each case the relevant parameters have been increased or decreased by 25% except in the case of survival without transplant where a 50% increase or decrease is applied to reflect greater uncertainty of these estimates. Green cells indicate greater cost-effectiveness, and red cells indicate low cost-effectiveness.

**Table 4.**
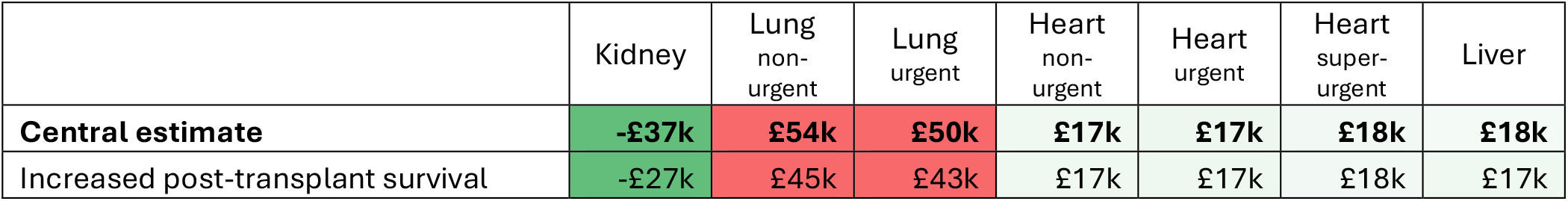
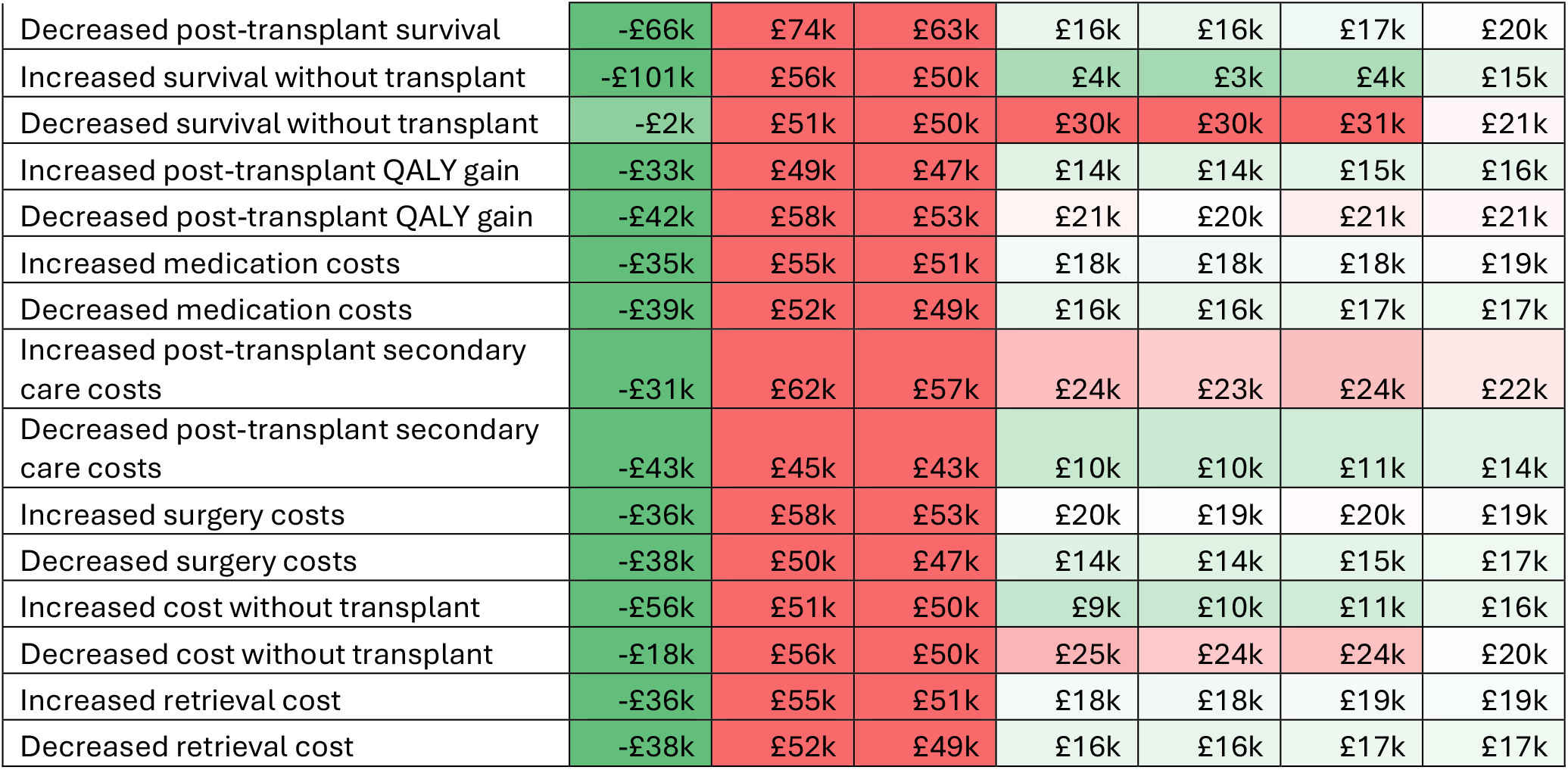
Sensitivity analysis of cost-per-QALY-gained.

|  | Kidney | Lung non-urgent | Lung urgent | Heart non-urgent | Heart urgent | Heart super-urgent | Liver |
| --- | --- | --- | --- | --- | --- | --- | --- |
| **Central estimate** | **-£37k** | **£54k** | **£50k** | **£17k** | **£17k** | **£18k** | **£18k** |
| Increased post-transplant survival | -£27k | £45k | £43k | £17k | £17k | £18k | £17k |
| Decreased post-transplant survival | -£66k | £74k | £63k | £16k | £16k | £17k | £20k |
| Increased survival without transplant | -£101k | £56k | £50k | £4k | £3k | £4k | £15k |
| Decreased survival without transplant | -£2k | £51k | £50k | £30k | £30k | £31k | £21k |
| Increased post-transplant QALY gain | -£33k | £49k | £47k | £14k | £14k | £15k | £16k |
| Decreased post-transplant QALY gain | -£42k | £58k | £53k | £21k | £20k | £21k | £21k |
| Increased medication costs | -£35k | £55k | £51k | £18k | £18k | £18k | £19k |
| Decreased medication costs | -£39k | £52k | £49k | £16k | £16k | £17k | £17k |
| Increased post-transplant secondary care costs | -£31k | £62k | £57k | £24k | £23k | £24k | £22k |
| Decreased post-transplant secondary care costs | -£43k | £45k | £43k | £10k | £10k | £11k | £14k |
| Increased surgery costs | -£36k | £58k | £53k | £20k | £19k | £20k | £19k |
| Decreased surgery costs | -£38k | £50k | £47k | £14k | £14k | £15k | £17k |
| Increased cost without transplant | -£56k | £51k | £50k | £9k | £10k | £11k | £16k |
| Decreased cost without transplant | -£18k | £56k | £50k | £25k | £24k | £24k | £20k |
| Increased retrieval cost | -£36k | £55k | £51k | £18k | £18k | £19k | £19k |
| Decreased retrieval cost | -£38k | £52k | £49k | £16k | £16k | £17k | £17k |

Under all sensitivity analyses kidney transplants are found to be cost saving whilst lung transplants are consistently the least cost-effective. The cost-effectiveness of heart transplants is highly sensitive to assumptions on survival and costs without a transplant. The cost-effectiveness of liver transplants appears to be comparatively stable with respect to any one change.

For most organs, survival rates are the most significant parameter along with costs without a transplant. For lung transplants, as survival without a transplant is very poor, the most significant parameter is post-transplant survival. Post-transplant secondary care costs and annual costs without transplant were also significant parameters for most transplants.

Table 5 shows the change from the central estimate of each sensitivity analysis with green cells indicating an increase in cost-effectiveness and red cells indicating a decrease.

**Table 5.**
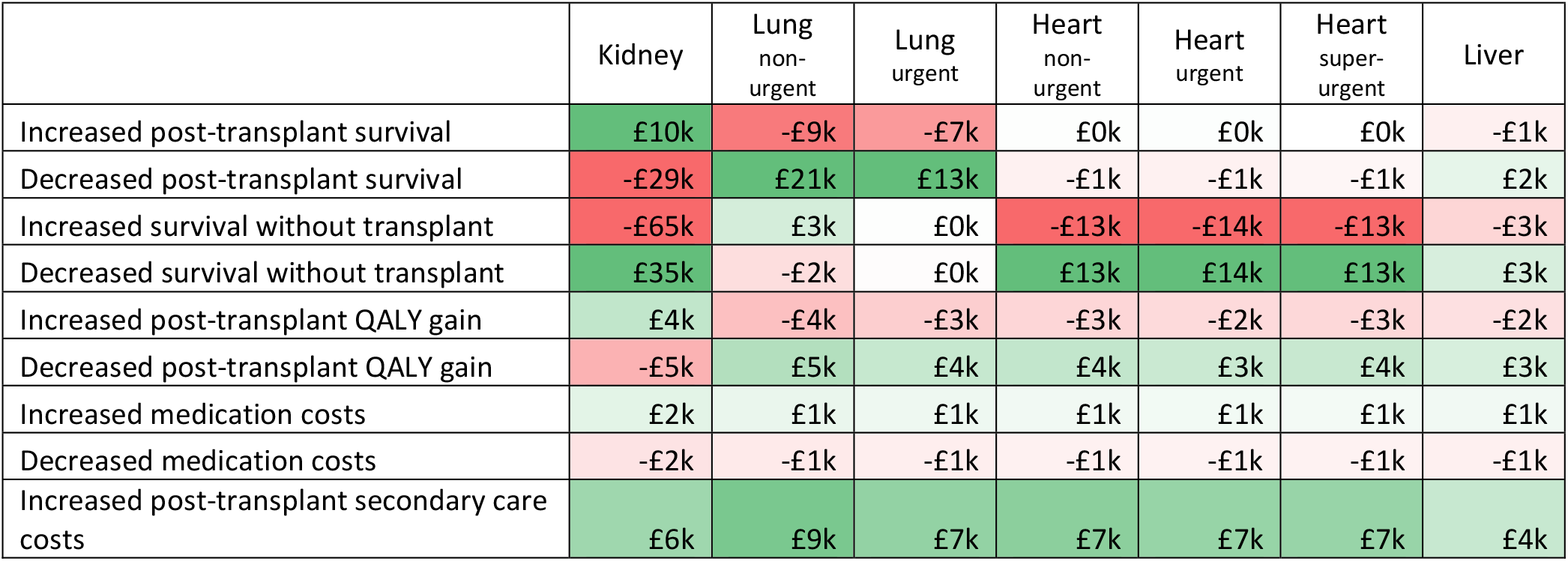
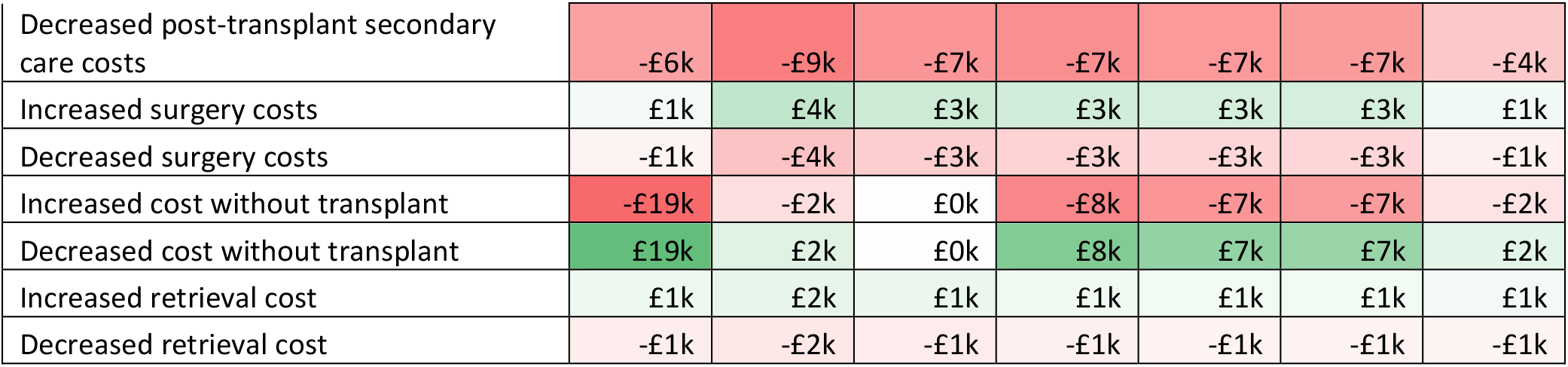
Sensitivity analysis: change from baseline.

## Conclusions

### The value of transplants to people and the healthcare system

The headline results of this analysis make it clear that kidney transplants are likely to be cost-saving interventions. This aligns with other studies such as [19-21].

Making more kidneys available for transplant by either increasing donation or ensuring more organs are of high enough quality to be transplanted could therefore save the NHS a significant sum in averted dialysis costs, whilst improving and extending lives. Furthermore, the negative impact of dialysis on the environment is well documented, e.g. [22] and kidney transplantation thus makes a positive contribution to the NHS Net Zero strategy.

Conclusions for other transplants will depend on the amount that is considered to be worth spending for one QALY. HM Treasury’s Green book gives a monetary willingness to pay value of £70,000 per QALY in 2020/21 prices [9]. At this level, transplants of all four organs considered in this paper are cost-effective. However, the Department of Health and Social Care (DHSC) typically uses a figure of £15k per QALY to represent the opportunity cost of spending in the NHS. This is based on analysis by Claxton et al that recommended a threshold of £12,936 per QALY [23]. To account for inflation and for simplicity a figure of £15,000 is often used. At this level lung transplants would not be cost-effective and both heart and liver transplants would be just outside the cost-effectiveness threshold in our central estimate though would be cost-effective in many of our sensitivity scenarios and would be cost-effective if retrieval is considered a sunk cost.

The National Institute for Health and Care Excellence (NICE) considers medicines to be cost effective if they cost less than £20-£30k per QALY [24]. It has recently been announced that this threshold will be raised to £25-£35k per QALY from April 2026. At this level kidney, heart, and liver transplants would all be considered cost-effective though lung transplants would not.

It is worth noting that the sensitivity analysis for survival without a transplant identifies a potentially counterintuitive result. Namely that, except for lungs, transplants are more cost-effective if survival without them is better. This is particularly noticeable for heart transplants. If a patient would have had very poor survival without a transplant, then a transplant could increase their life-expectancy significantly gaining many QALYs. However, poor survival without a transplant is also cheaper, meaning a transplant does not avert as many costs.

It also suggests that if more successful ways of keeping people alive without a transplant are developed, unless those approaches are cheaper than current options, transplants may become even more cost-effective over time.

### Strengths and limitations of this work

A key strength in this work is that the same approach was used across four organ types leading to a fair comparative estimate of cost-effectiveness. This also allows for an assessment of the total benefit that can be attributed to a single donor who may donate multiple organs.

Our approach considers whole health system costs for the full lifetime of a patient to account fully for impacts on life-expectancy and ongoing costs of treatment.

The sources used for the health-related quality of life utilities weights are over twenty years old highlighting the need for more research to quantify the impact of transplants on quality of life.

Several key simplifications were made in both methodology and scope. The approach in effect considered an “average patient” but in practise the variation between pathways and outcomes for different patients can be significant. The analysis also focused solely on adult transplants using deceased donor organs. Future work could look in more detail at different patient pathways and expand the scope to include paediatric transplants, living donor transplants for kidney or liver lobe, and other types of transplants such as pancreas or multi-organ transplants.

### Wider benefits of transplants

This work focussed on impacts to patients’ health and the healthcare system. It did not look at wider effects such as impacts on patients’ employment, benefits payments, or the impacts on the families of patients.

Many transplant recipients are of working age and following transplant may return to sufficient good health to return to work. A systematic review by D’Egidio et al found that an average of 39.4% of kidney transplant recipients return to work within a year [25]. For liver and heart transplants there is some evidence that fewer people are in work after transplant than before transplant however it is unclear whether those who continued to work post-transplant would have been able to do so without it [26 & 27]. A recent paper showed that an increase from 19.5% working at the time of transplant to 35.5% working after transplant for those receiving a lung transplant in Australia [28].

More evidence would be needed to be able to quantify the economic benefits of transplants in the UK with regards to employment.

## Data Availability

All data produced in the present study are available upon reasonable request to the authors

